# Protocol for a Systematic Review to validate or not, the philosophic explanation under the mechanism of action of the Tension Band Wiring to estabilizase an Olecranon fracture

**DOI:** 10.1101/2022.01.18.22269464

**Authors:** Sergio Barcia, Jorge Boretto

## Abstract

Olecranon fractures in young people are commonly treated surgically. When a transverse pattern is presented the most used method to fix the fracture is the Tension Band Wiring. The AO Foundation proposed a theory for the mechanism of action of this device. It could transform the tensile forces on the posterior side into compression forces on the joint side during flexion of the elbow. Our objective is to systematically review the available literature in order to confirm or not this hypothesis.

We will include biomedical research studies evaluating Tension Band Wiring function. We are planning to search on Medline, Embase and other sources including grey literature. The studies will be selected and the data extracted. Results will include a descriptive analysis and a meta-analysis to compare the observed compression force before and after the flexion of the elbow.

Our findings will be discussed having in mind the potentially clinical implications of results.

## Introduction

Fracture treatment faced a revolution that begun when surgery was safe after the introduction of aseptic techniques. It was followed by the concept of stable fixation using plates and screws and by the development of biologically inert material to avoid corrosion. It was Robert Danis who considered a callus formation as a pathological structure to be avoided by inter-fragmentary compression in 1949. In 1950 four Swiss surgeons joined to study and teach modern osteosynthesis techniques forming the AO group. (Augat P. and von Ruden Ch., 2018).

The AO Foundation is now a core institution that helped directly or indirectly to save lives and improved patients outcomes after fracture treatment around the world. Tension Band Wiring was introduced by Weber and Vasey in 1963 for the treatment of Olecranon fractures and spread by Muller and the AO school since 1970 (Sangines F.,1988). While plates, screws and nails had evolved incorporating new designs and new technologies, the frequently used Tension Band Wiring device remains as the same device it was early designed. There is a current description of it at the Surgery Reference guidelines of the AO Foundation web site (Kloen P. and Ring P., 2018).

The anual incidence reported for proximal ulna fracture is 15 per 100.000 population. It represented the 21% of the proximal forearm fractures, and the 1,1% of the totality of fractures recorded prospectively in a trauma centre in Edinburgh in a twelve month period (2007-2008). Olecranon fractures represented the 82% with an incidence of 12 per 100.000 population. The most frequent type of fracture was a simple displaced one (73,5%). And the most frequent way of treatment was surgical reduction and fixation with a Tension Band Wiring, in 41 out of 47 cases (Duckworth A., 2012).

There are four principles for surgical fracture fixation: anatomical reduction, blood supply preservation, stable stabilisation (absolute or relative) and early and safe mobilisation. (Bizzarro J. and Regazzoni, P., access on line 16/8/21) For an Olecranon simple fracture, without conminution neither joint instability, there is not discussion about the high possibility to achieve the anatomical reduction in an atraumatic way, with a soft tissue and bone gently handling in order to preserve the blood supply. But some question should be answer to explain what means safe early mobilisation and what kind of stability provides the most used device for fracture fixation in this kind of fracture.

An articular fracture is the main indication for an absolute stability fracture fixation. AO defined absolute stability as a fixation method with: inter-fragmentary compression, no movement at the fracture site and no callus formation. The AO considered there are three devices to achieve the inter-fragmentary compression: the Lag Screw, the Compression Plate, and “ the Tension Band”.(Bizarro J. and Regazzoni P., access on line 16/8/21)

In textual words The AO experts said : “The philosophy behind the tension band for the Olecranon is that it converts tensile forces on the posterior side of the olecranon into compression forces at the joint line during flexion. The fixation is simple and inexpensive and works well if executed properly” (Kloen P. and Ring P., 2018). But, what does philosophy means? The Oxford English Language dictionary provides the following definition for the noun “philosophy”: “a theory or attitude that acts as a guiding principle for behaviour”(Oxford Languages and Google, accessed on line 16/8/21). A theory or actitud is not enough for a recommendation about a surgical procedure. Instead, an evidence based approach is required to support a therapeutic recommendation in contemporary times.

The primary aim of this Systematic Review is to find evidence in biomechanical research to support or reject the “Tension Band Wiring theory”. Our secondary aim is to define the type of stability the TBW provides to the fracture or the osteotomy site in the Olecranon. To this end the Systematic Review will answer the following questions:

1. Does the TBW provides inter fragmentary compression?
2. Does the compression forces increase at the joint line during flexion of the elbow joint?
3. What is the difference of compression forces in the static fase, between TBW and other devices?
4. What is the maximal load resistance for tension forces of the TBW?
5. What is the difference in tension resistance, between TBW and other devices?
6. What are the number of cycles necessary for TBW failure?
7. What is the difference in number of cycles to failure, between TBW and other implants?
8. What is the maximal torsional forces that TBW is able to resist?
9. What is the difference in torsional forces resistance, between TBW an other devices?
10. What are the mechanisms of failure in TBW?

## Methods

### Eligibility

We will include the following studies:

- RCT and observational studies
- Performed on synthetic or cadaveric bones.
- They must be biomechanical research on Olecranon fractures or osteotomies
- They will study the TBW alone or comparing or describing different osteosynthesis devices.
- They need to include objective measurement of compression forces, tension forces resistance, torsional forces resistance, or/and cycles to load resistance.
- There will be no language restriction. Spanish and English reports will be read by the main authors and one reviewer also reads French language. Studies reported in other idioms will be translated to Spanish using Google Translate.
- Without restriction for year of publication
- Clinical studies reporting clinical outcomes, like complications or functional scores outcomes, on patients will be particularly excluded.
- Experiments performed on animals will also be excluded.

## Search

Literature search strategy will include MeSH terms (medical subject heading) and text words related to biomechanics, Olecranon and Tension Band Wiring.

We will search Medlline database on Pubmed library, EMBASE on Ovid interface, Google scholar, Web of science and ILIACS. We also will search on References from systematics reviews on Olecranon fractures and on Systematics reviews about biomechanics. We will look for citations on included studies and on those selected ones to discuss later on Discussion section. Those papers available on “Related articles” on Pubmed will be also added to the search. We will seek for on web sites available in Google dedicated to Osteosynthesis, like AO Foundation and Medartis. The search will be completed by including protocols on PROSPERO and ClinicalTrials. gov.

The following one, will be the search strategy for Medlinine database including MeSH terms and text words in title and abstracts: *Biomechanics OR Biomechanics OR Biomechanics Phenomena OR experiment* OR synthetic bone OR cadaveric bone AND olecranon OR olecranon process OR Tension Band OR Cerclage NOT humans NOT animals*

## Selection

Two reviewers independently will select publications after carefully screening of titles and abstracts guided by inclusion criteria. This initial selection will lead to the review of the full text publication. Any disagreement between authors will be solved by discussion and consensus. If necessary, the studies authors will be contacted for more details to decide its inclusion. Neither of the review authors will be blinded to original authors and institutions during the search and selection steps.

## Data extraction

Two authors will independently extract the data from selected publications. A pilot exercise is planned for training in order to reduce mistakes during this important step. The extraction form will be written on an Excel format page (or a similar one).

We will extract the following information:

- Publication and the original authors: title, name, year of publication, institution and country;
- The type of experiment: cadaver bones or synthetic bones, number, type of osteotomy;
- Setting technique: cerclage configuration, comparison (if exist) and testing mechanism.
- Outcome measurements: failure mechanism, tension resistance, compression, cycles to load resistance, gap distance, torsion resistance
- Comments from authors

The primary outcome will be a quantitative variable to express compression difference at the simulated fracture, between basal set up and after flexion of the elbow by simulating active or passive motion of a joint. Secondary outcomes will express mechanism of failure, time or forces to failure and basal compression (static).

## Risk of Bias for individual studies

We expect to include a wide range of study designs, from descriptive ones to randomised and non randomised comparative studies. To evaluate the internal validity on every included study, we specially will asses the experimental setting description and the quality of artificial or human bones. In comparative studies we will explore how authors intended to avoid selection bias. Performance bias will be accessed in order to confirm if specimens and fixation techniques were performed equally in all the sample. Outcome measurement and reporting will be the most relevant source of bias. We will need to know who did the evaluation and how the outcome was measured. If any piece of experiment was excluded we will find the reason for that decision. Any conflict of interest will be recorded. The bias assessment tools from CEBM will be used to guide this step in the review (https://www.cebm.net). Two review author will independently asses risk of bias, and any disagreement will be addressed by discussion and consensus. Eventually, an opinion from another author will be required if it is necessary.

## Data synthesis

### Descriptive

A Table to describe the characteristics of the experiments reported will be obtained. It will contained: the name of studies selected, year of publication, type of bones used (cadaveric or synthetic), setting of the experiment, type of TBW (bi-cortical K-wire, wire diameter, number of knots, screws enter k wires, sutures enter wires, etc) sample size, specimen lost, main results, and authors conclusion.

### Primary analysis

When two comparative cohorts exists, the single cohort where tension band was studied will be selected first for the primary analysis. If different units of measurement are used to quantify the compression force on site of osteotomy, a standardisation procedure will be needed to allow for a data synthesis analysis.

A Meta-analysis to summarise the difference of compression forces before and after elbow flexion with TBW will be the main analysis. The result will be a continuos variable, so a standard mean difference with 95% CI will be obtained as the main outcome. It will be valid to answer wether the tension band theory is validated or not. A subgroup analysis considering different designs of the TWB will be run.

Secondary analysis will be the following ones:

- A Meta-analysis to summarise the static compression force of TBW, using inverse variance method.
- A Meta-analysis to compare the static compression force between TBW and other devices, with subgroup analysis considering different devices.
- A Meta-analysis to summarise the maximal tension load resistance of TBW, using inverse variance method.
- A Meta-analysis comparing the maximal tension load resistance between devices, with subgroup analysis considering different devices,
- A meta-analysis to summarise the maximal cycle resistance of TBW, with inverse variance method.
- A Meta-analysis comparing maximal cycle resistance between devices, with subgroup analysis considering different devices.
- A Meta-analysis to summarise the maximal torsional forces resistance for TBW
- A Meta-analysis comparing maximal torsional forces resistance between devices, with a subgroup analysis considering different devices

Heterogeneity will be explored by analysis the settings, types of specimens and techniques of experimentation. I squared (50% or more will be considered as substancial heterogeneity) and Chi squared test (significant level 0,1) will be valid to measure statistical heterogeneity. SPSS software will be used to all of the statistical analysis, Random effect meta-analysis will be chose if heterogeneity is confirmed, otherwise a Fixed effect method will be selected as the primary method. However, both type of analysis will be run considering the possible effect of small sample size on result.

A subgroup analysis by type of material (cadaveric or synthetic), and technique used for tension band configuration will be the way to explore the robustness of the result. A sensitivity analysis including only studies with lower risk of bias is also planned for that purpose.

A narrative synthesis will be the main result if data cannot be quantitatively summarised and it will be also important as complement for a quantitative result. The table with the selected studies their methods, results and conclusions will be useful to guide and complement the narrative comparison.

Funnel Plot will be used to identify publication bias. We will search for protocols of the included studies in order to detect if any incongruence exists on the outcome or the technique designed for the experiments.

The quality of the evidence will we decided with the GRADE approach. It will be important to support the main conclusion.

## Data Availability

This is a Protocol for a Systematic Review. There is not any new data yet.

